# Increased variance in second electrode accuracy during deep brain stimulation and its relationship to pneumocephalus, brain shift, and clinical outcomes: a retrospective cohort study

**DOI:** 10.1101/2022.03.06.22271981

**Authors:** MG Hart, M Posa, PC Buttery, RC Morris

## Abstract

**Introduction:** Accurate placement of deep brain stimulation electrodes within the intended target is believed to be a key variable related to outcomes. However, methods to verify electrode location are not universally established.

**Research Question:** The aim of this study was to determine the feasibility of post-op lead localisation in clinical practice and its utility to audit our own DBS accuracy.

**Material and Methods:** A retrospective cohort study was performed of a consecutive series of patients with Parkinson’s disease who underwent deep brain stimulation of either the globus pallidus internus (GPi) or subthalamic nucleus (STN) between 2016 and 2019. Image processing was performed using the Lead-DBS toolbox. Institutional ethical approval was granted as a review of service.

**Results:** In total 38 participants met the inclusion criteria. Electrode localisation was completed in 79%. Clinical outcomes included improvement in UPDRS III of 46% and PDQ39 of 32%. Overall electrode accuracy was 0.22 +/- 0.4mm for all electrodes to the main nucleus with 9 (12%) outliers but only 3 (4%) electrodes out with 2mm from the intended target. Accuracy was worse for the second electrode implanted and in the GPi but was not affected by pneumocephalus or brain shift. Neither clinical outcomes nor the volume of activated tissue was affected by electrode accuracy.

**Discussion and Conclusions:** A neuroimaging approach to electrode localisation allows qualitative appraisal of targeting accuracy and is feasible with routine clinical data. Such methods are complimentary to traditional co-ordinate approaches and lend themselves to developing large, collaborative, and quantitative projects.

**HIGHLIGHTS:**

- Overall electrode accuracy was 0.22 +/- 0.4mm with only 3 (4%) electrodes out with 2mm from the intended target
- Accuracy was significantly worse in the GPi versus the STN and on the second side implanted
- Inaccuracy occurred in the X (lateral) plane but was not related to pneumocephalus or brain shift

## INTRODUCTION

Deep brain stimulation for Parkinson’s disease is an effective treatment supported by evidence from randomised controlled trials[1–5] and is now established in routine clinical practice internationally. Despite the ubiquity and success of the procedure, methods for appraisal of electrode accuracy – a key surgical outcome variable[6–9] – have not yet been formalised. Failure to appreciate electrode accuracy may lead to excessive revision rates, unsatisfactory clinical outcomes, and an inability to effectively appraise surgical results.

Placement of electrodes has historically relied on microelectrode recordings (MER) and intraoperative macrostimulation during awake surgery[10]. However, the field of deep brain stimulation has evolved towards image-guided surgery under general anaesthesia without MER for reasons of efficiency, patient comfort, and safety[11–14]. Methods for appraising electrode accuracy are therefore necessarily based on imaging, and while commercial and open-source software is available for this purpose[15–21], reports of applications of these methods in routine clinical practice are few.

Our aim was to test the applicability of image-based electrode localisation, specifically using the open-source Lead-DBS toolbox[17,21], in routine clinical practice. In particular, we wished to test the ability to interrogate the accuracy of our own deep brain stimulation practice for Parkinson’s disease using a direct targeting, MRI guided, and CT verified technique under general anaesthesia. As a proxy for the accuracy of electrode localisation, we hypothesised that the volume of the target nucleus that was stimulated would correspond most strongly with motor outcomes. Furthermore, we hypothesised that inaccuracy would be related to well-known variables, namely pneumocephalus and intraoperative brain shift.

## MATERIALS AND METHODS

### Patients

A retrospective cohort study was performed of a consecutive series of patients with Parkinson’s disease who underwent deep brain stimulation of either the GPi or STN between 2016 and 2019. Patient selection for deep brain stimulation was performed in a multi-disciplinary setting according to national commissioning criteria[22,23]. Targeting of the subthalamic nucleus (STN) was favoured when levodopa equivalent daily dose (LDD) reduction was the aim, while targeting of the globus pallidus internus (GPi) was favoured if there were concerns regarding cognitive decline, tolerance of programming, or if LDD was not a priority. Baseline patient details are presented in table 1. Institutional ethical approval was granted as a review of service study.

### Surgical Procedure

Surgery was performed as a single stage procedure under general anaesthesia with implantation of electrodes manufactured by: St Jude/Abbott (Abbott Laboratories, Lake Bluff, Illinois, USA: 6147 non-directional electrode with Libra PC system, or 6170 directional electrode with Infinity system), Medtronic (Medtronic Inc, Dublin, Eire: 3389 electrode with Activa PC system), or; Boston (Boston Scientific, Marlborough, Mass, USA: Cartesia directional electrode and Vercise PC or Gevia system). Planning was performed using StealthStation® S7™ (Medtronic Inc, Dublin, Eire) FrameLink® software. Direct targeting was performed based on pre-operative 3 Tesla MRI data (magnetization-prepared rapid-acquisition gradient-echo (MPRAGE) volumetric STEALTH sequences, proton density (PD) sequences for the GPi, and susceptibility-weighted imaging (SWI) sequences for the STN. A Leksell frame (Elekta AB, Stockholm, Sweden) was used in combination with pre-operative and post-operative volumetric CT imaging for trajectory planning and verification, respectively. If post-operative CT imaging demonstrated satisfactory appearances the implantable pulse generator was placed during the same general anaesthetic. In all cases the left hemisphere was implanted first as it typically reflected represented the dominant hemisphere.

### Image Registration, Electrode Reconstruction, and Calculation of Volume of Activated Tissue (VAT)

Image processing was performed using the Lead-DBS toolbox [17,21] (figure 1A). Registration of the post-operative CT image to the standard space template of the Montreal Neurological Institute (MNI152) was performed using Advanced Normalization Tools (ANTs)[24,25]. Specifically, post-operative CT images were linearly registered to the pre-operative T1-weighted MPRAGE image which in turn was registered to the ICBM152 2009b template with non-linear diffeomorphic warping. Brain shift correction of subcortical structures was performed using linear registration of an additional subcortical mask[26].

**Figure 1:**
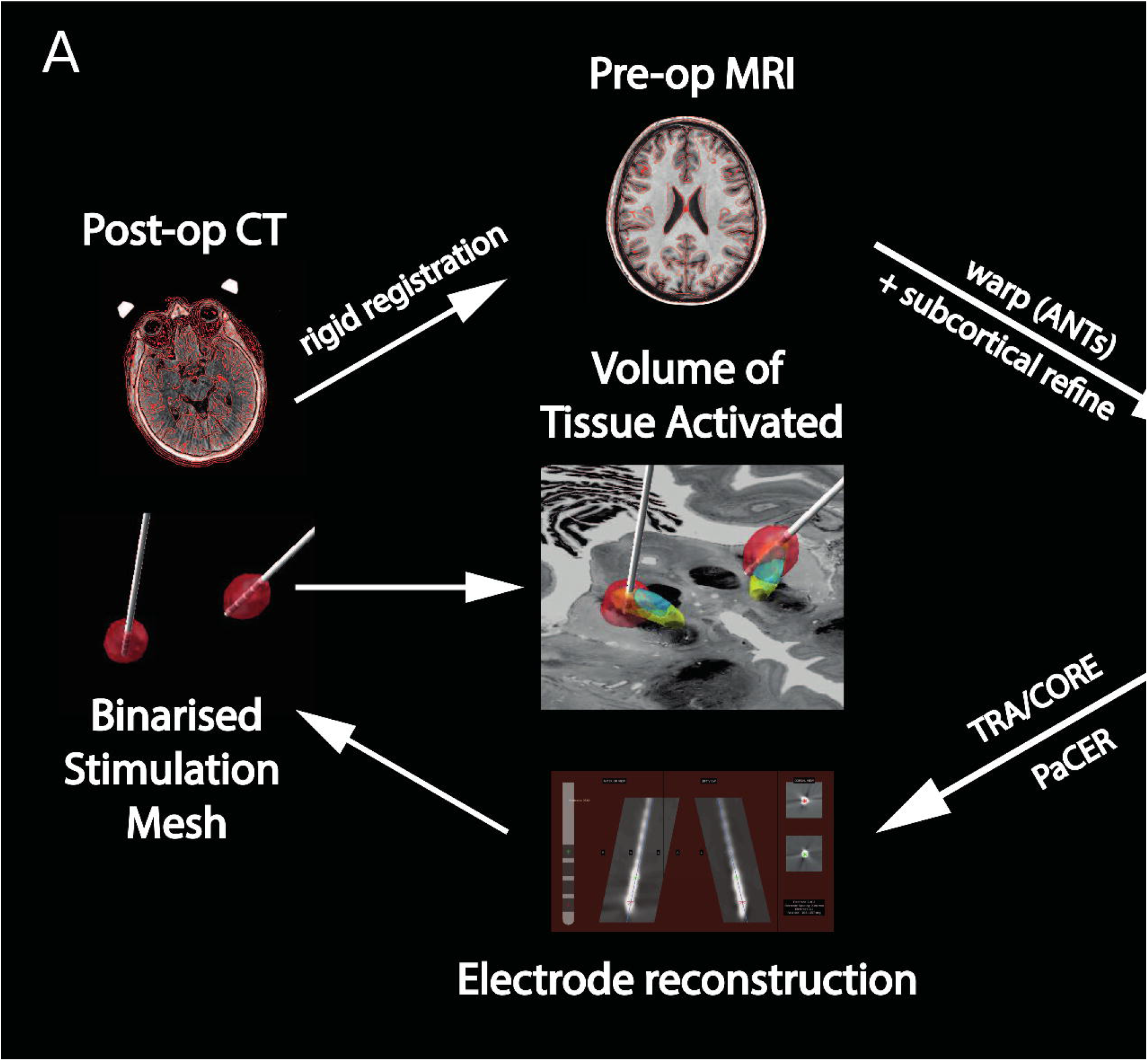
Image Processing Pipeline. A: Post-operative CT scans were linearly registered to the pre-operative MRI which in turn was registered to the standard space of the Montreal Neurological Institute using non-linear diffeomorphic warping. Subsequently, subcortical structures were subject to an additional rigid registration to account for brain shift (subcortical refine). Both transforms were combined and applied. Electrode reconstruction was performed using either PaCER for non-directional leads or TRA/CORE for directional leads. Subsequently a Volume of Activated Tissue (VAT) was generated as a mesh and displayed on a template atlas. B: electrode distances were defined according to distance from nucleus border (right) for analysis of accuracy, or distance to either the main nucleus (grey asterisk) or motor subnucleus (white asterisk) when analysing XYZ variance.

Electrodes were reconstructed based on the post-operative CT images using a combination of PaCER[20] algorithm or TRA/CORE[17,21] approach depending on individual requirements. Subsequently manual refinement was performed to align the electrode model with the imaging artefact on the corresponding CT image.

Volume of Activated Tissue (VAT) reconstructions were performed using a finite element model implemented in Lead-DBS[17,21]. A tetrahedral mesh was constructed based on a four-compartment model comprising grey matter, white matter, and electrode (conducting and non-conducing) components. Conductivity values were set according to standard parameters then the VAT was binarised at a threshold of 0.2 Volts/mm. Finally, electrode locations and VATs were visualised on the DISTAL atlas[27].

### Definition of Accuracy

Electrode accuracy was defined as the distance between any electrode contact and the boundary of the target region (either main nucleus or motor subnucleus) (figure 1B). This was defined in both the 2D plane for target plots and in terms of 3D Euclidean distance. Electrode variance was defined as the distance in 3D Euclidean co-ordinates from the centre of gravity of the target in question (either the main nucleus of motor subnucleus).

### Computation of Brain shift and Pneumocephalus

Brain shift is believed to occur after durotomy and results in a complex and likely nonlinear displacement of the brain that is poorly defined. This can result in significant difficulties in registration between images as nonlinear transforms can impact on electrode localisation robustness. Subsequently, one of the most well-developed methods to account for this relies upon using layered linear transforms to subcortical regions of interest ([26]). We quantified this approach by summing the resultant transformation matrix. Additionally, we localised electrodes both with and without this approach to assess the difference it made to accuracy. Pneumocephalus is typically related to durotomy and believed to be associated with brain shift. We quantified the degree of pneumocephalus by using an MNI space brain mask which defined pneumocephalus as the difference between this as the expected brain volume and the actual extracted brain volume (figure 1C).

### Outcome Assessment

Programming commenced at approximately 6 weeks following surgery and was led by a consultant neurologist with a specialist interest in movement disorders in combination with a specialist nurse. Initial programming commenced with a pulse width of 60μs and a frequency of 130Hz. Outcome variables were recorded by a consultant neurologist in combination with a specialist nurse at latest follow-up (and at least 12 months following surgery). Outcome measures included: body weight (in kilograms); UPDRS 3; UPDRS 4; levodopa equivalent daily dose (LEDD); and PDQ39.

### Statistical Analysis

Groups were analysed according to hemisphere (with the second electrode to be implanted being in the right hemisphere), nucleus (GPi or STN), and target (main nucleus or motor subnucleus). Distances from the electrode to, and overlap of the VAT with, the corresponding nucleus and specific component of the nucleus were calculated. All values are expressed as mean +/- 2 standard deviations (SD). Raincloud plots were generated to display raw data, box plots, and half-violin plots of the data distribution ([28]).

Differences in continuous variables were performed with paired t-tests (dual groups) or Analysis of Variance (ANOVA, multiple groups). Statistical dependencies between continuous variables were analysed with Pearson’s correlation. A general linear model was fit on clinical outcomes, electrode accuracy, and VAT’s. Significant was set at p<0.05 with corrections for multiple comparisons using the Bonferroni method. All analyses were performed in MATLAB (version 9.7.0 (R2021a), Natick, Massachusetts: The MathWorks Inc.) using open-source code (https://github.com/jazzmanmike/DBS/).

## RESULTS

### Cohort Characteristics

In total 38 participants met the inclusion criteria. Baseline clinical and demographic data of the cohort are presented in Table 1. The only clinical adverse events were a device infection that required system explantation and tethering of an implantable pulse generator managed with revision surgery.

### Image Processing

Overall, 30 out of 38 (79%) participants successfully completed the image processing pipeline (supplementary figure 1). Reasons for exclusion from analysis included registration failure (post-op CT to pre-op MRI, 6 participants) and failure of VAT construction (2 participants). Correction for brain shift with subcortical refine methodology was utilised in all cases. Electrode reconstructions were performed primarily with PaCER, or TRA/CORE if this was not possible (15 participants each).

### Clinical Outcomes

Changes in clinical outcomes post-operatively are presented in figure 2 and table 2. In the overall cohort, statistically significant improvements were demonstrated for UPDRS 3, UPDRS 4, LEDD, and PDQ39. Stimulation of the STN compared with the GPi resulted in a significantly greater reduction in LEDD (55% decrease versus 2% increase respectively, p<0.001) but otherwise there were no differences in outcomes. Finally, all the described clinical outcomes were independent and did not demonstrate significant covariance (supplementary figure 2: maximal R^2^ 0.13, p = 0.12).

**Figure 2:**
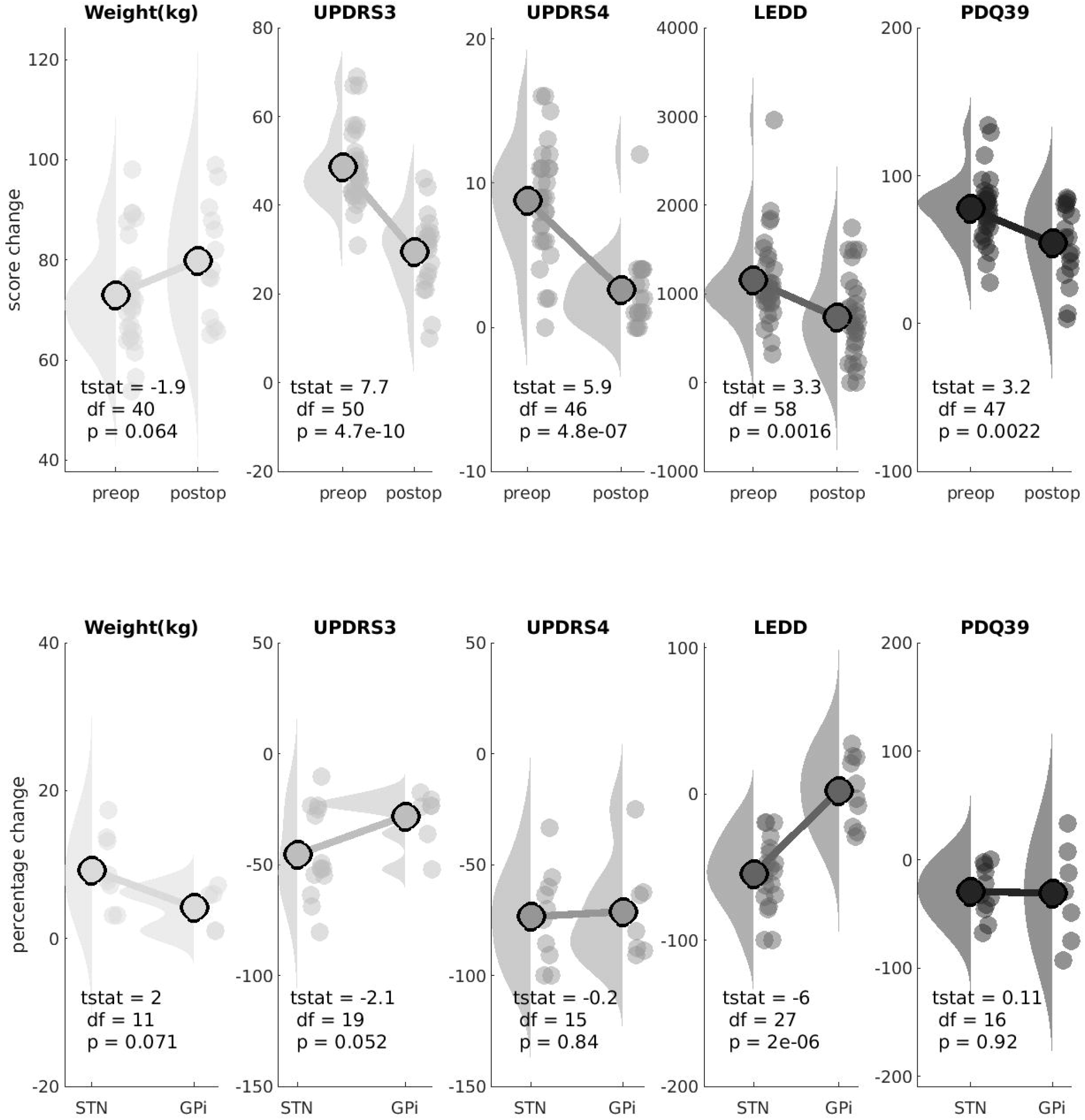
Clinical Outcomes. Raincloud plots of changes in the five clinical outcome variables. Upper rows (green) are pre-operative values, lower rows (orange) are post-operative scores. All changes are as percentages. tstat = paired t-test value. Raincloud plots display the raw data, box plots, and half-violin plots of the data distribution ([28]).

### Electrode Accuracy

Overall electrode accuracy was 0.22 +/- 0.4mm for all electrodes to the main nucleus with 9 (12%) outliers but only 3 (4%) electrodes out with 2mm from the intended target (figures 3 and 4). For GPi stimulation, electrodes were a mean of 0.26 +/- 0.43mm from the main nucleus, and 0.80 +/- 0.97mm from the GPi motor nucleus. Overall, 17 out of 20 electrodes were within 2mm of the main nucleus. For STN stimulation, electrodes were a mean of 0.14mm (SD 0.28) from the main nucleus, and 0.27mm (SD 0.52) from the STN motor nucleus. Overall, all 40 electrodes were within 2mm of the main nucleus.

**Figure 3:**
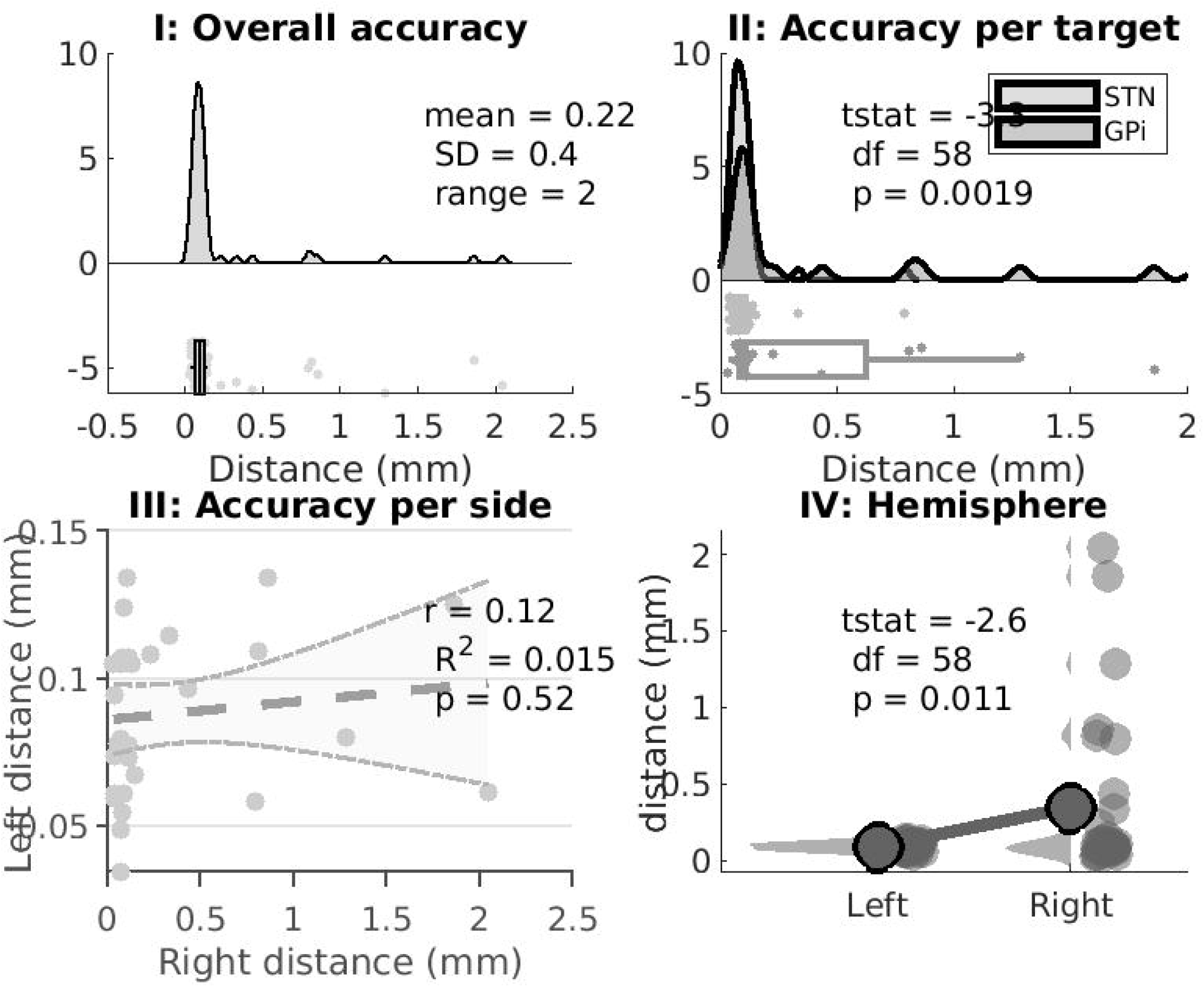
Electrode Accuracy Summary. I: Overall accuracy for all electrodes for both targets (GPi and STN) and hemispheres (60 electrodes). In total 9 outliers were identified and only 3 electrodes out with 2mm from the intended target. II: Electrodes implanted in the GPi had lower accuracy than those implanted in the STN. III: Electrode accuracy did not systemically vary between sides (i.e. it was not the case that low accuracy in one hemisphere was associated with low in the other hemisphere, related to for example a shared methodological step). IV: Electrode accuracy varied between hemispheres with reduced accuracy in the right hemisphere which was the second side implanted.

**Figure 4:**
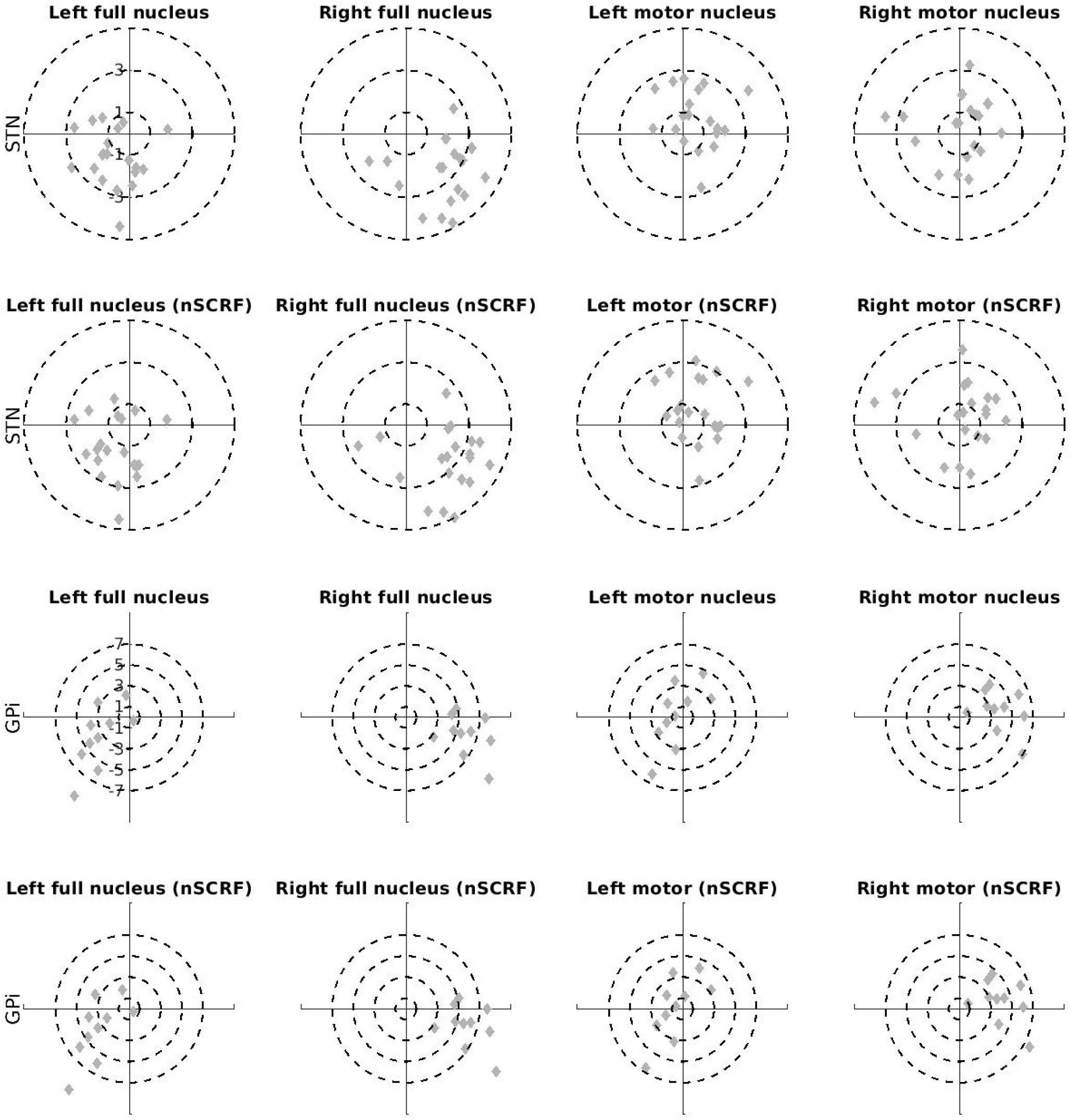
Group Electrode Location Visualisation. A: subthalamic nucleus electrode reconstructions B: corresponding subthalamic nucleus electrode contacts on the right (upper) and left (lower). C: globus pallidus internus electrode reconstructions D: corresponding globus pallidus interna electrode contacts on the right (upper) and left (lower). The background image for all figures is from the BigBrain project (https://bigbrain.loris.ca/main.php?) under CC-BY-NC-SA 4.0 license.

Nucleus (GPi or STN) affected accuracy with lower accuracy for electrodes in the GPi than the STN (0.43 +/- 0.62mm versus 0.11 +/- 0.12mm, p = 0.002). There was no systematic co-variance in accuracy between hemispheres (r = 0.12, R^2^ = 0.01, p = 0.52). However, the second electrode implanted (i.e. that in the right hemisphere) was less accurate than the first electrode implanted (0.09 +/- 0.03mm versus 0.34 +/- 0.53mm, p = 0.01).

To determine which clinical scenario (GPi versus STN, main nucleus versus motor subnucleus, right versus left side) accuracy is most likely to be affected, a systematic analysis is presented in supplementary figure 3. Concordant with the main findings above, variance in accuracy was most prominent in the right GPi (group F-stat = 9.68, df = 7, p < 0.01; GPi right tstat = -2.9, df = 18, p = 0.01). Accuracy did not vary depending on whether the target reference was chosen to be the main nucleus or motor sub-nucleus (tstat = -1.1, df = 118, p = 0.27).

### Systematic Variance in Accuracy

Target plots of accuracy per nucleus, hemisphere, and target are presented in figure 5 to determine if variation in accuracy occurred predominantly in any single Cartesian (XYZ) dimension. Variance occurred predominantly in the X-dimension in the right hemisphere and was consistent across nucleus (STN versus GPi) and target (overall nucleus versus motor sub-nucleus) (supplementary figure 4). Addition of subcortical refine methodology (as a proxy for brain shift) did not affect accuracy in any single dimension.

**Figure 5:**
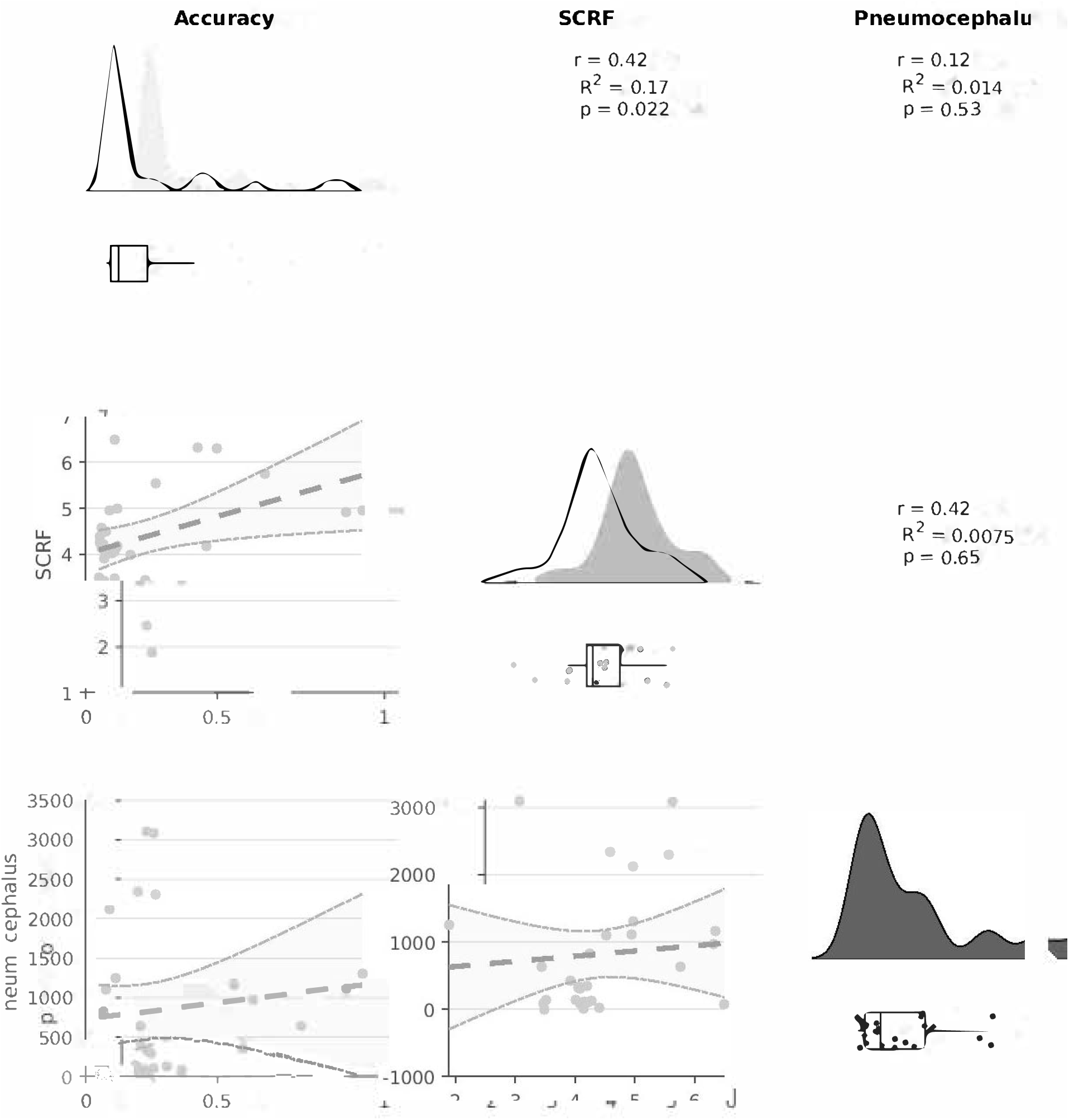
Target Plots of Electrode Location. Electrode co-ordinates are displayed in relation to the target centre of gravity (see figure 1B left). This allows systematic visual inspection of lateral (X) and anterior (Y) plane variance viz a viz accuracy and precision of group targeting. These plots are systematically arranged per target (STN: upper two rows, GPi: lower two rows), nucleus (main: left two columns, motor: right two columns), and hemisphere. Rows two and four remove the subcortical refine (nSCRF) that compensates for brain shift from the corresponding co-ordinates in the row above.

### Relationship of Accuracy to Pneumocephalus and Brain Shift

Analysis of factors that may impact upon accuracy, specifically pneumocephalus and brain shift, is presented in figure 6. Accuracy, in this instance defined as the mean accuracy across hemispheres, positively correlated with brain shift, as defined by the total adjustment performed by subcortical refine methodology, but this was not significant when corrected for multiple comparisons (r = 0.42, R^2^ = 0.17, p = 0.022). There was no correlation between accuracy and pneumocephalus, nor between brain shift and pneumocephalus.

**Figure 6:**
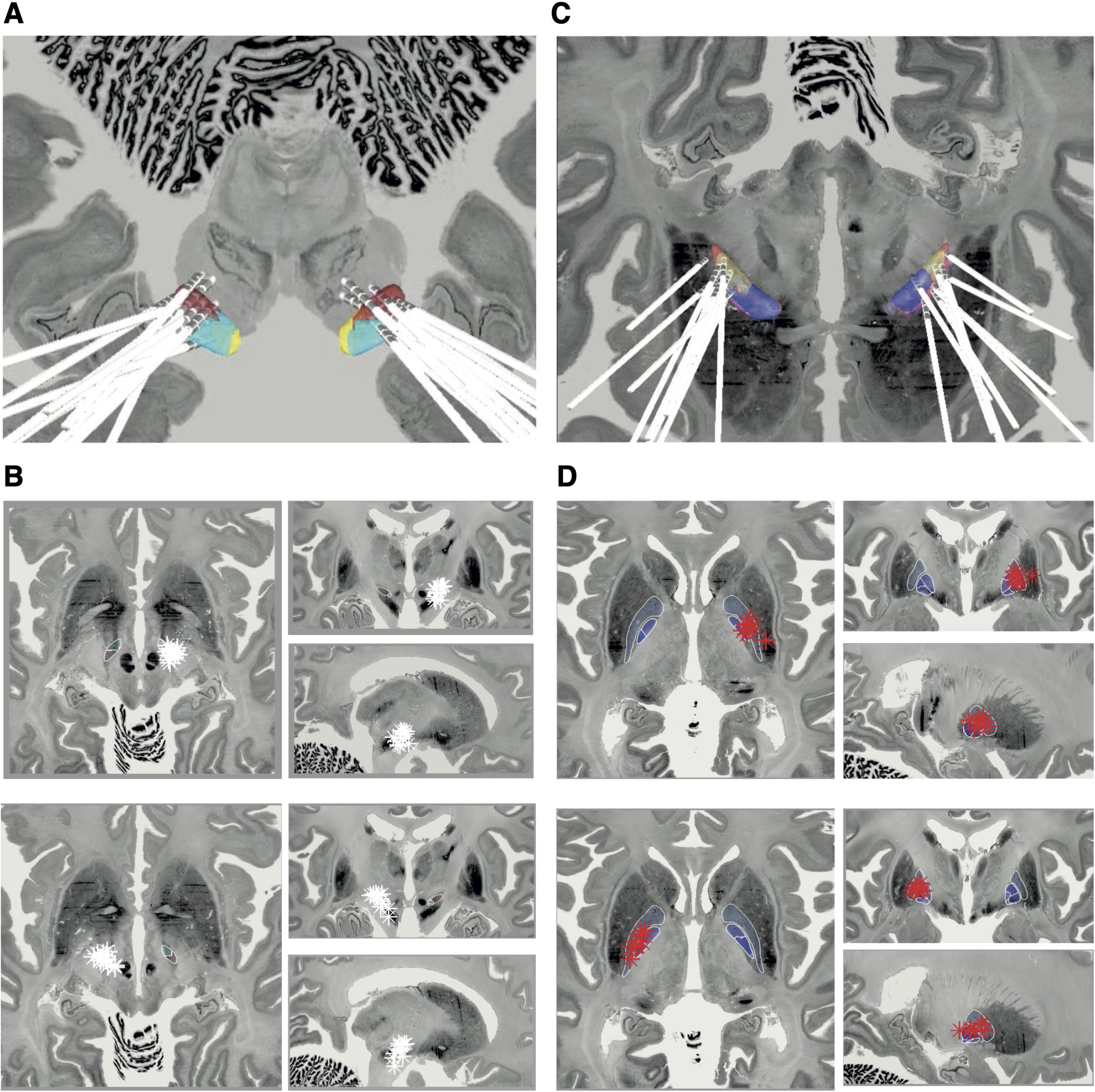
Accuracy, Pneumocephalus, and Brain Shift. Electrode accuracy and averaged across hemispheres. Brain shift is reflected in the summed total of the subcortical refine (SCRF) transformation matrix. Pneumocephalus is assessed using a standard space brain mask (figure 1C). Increased brain shift, reflected in higher SCRF values, is correlated with reduced accuracy. However, accuracy is not correlated with brain shift, and brain shift is not correlated with pneumocephalus.

### Accuracy and Clinical Outcomes

Finally, the relationship of accuracy to clinical outcomes is presented in supplementary figures 5 and 6. Note that for GPi stimulation there were insufficient outcome data available for analysis and these data were excluded, hence these analyses are exclusively for STN stimulation. The VAT in either the main nucleus or motor sub-nucleus did not correlate with clinical outcomes. Furthermore, overall electrode accuracy did not correlate with the VAT in either the main nucleus or the motor nucleus (r = -0.19, p = 0.43).

## DISCUSSION

In summary, we performed a systematic appraisal of DBS electrode accuracy using contemporary neuroimaging methods. Overall, accuracy was high. However, accuracy was lower in the GPi than STN, and for the second electrode implanted. This inaccuracy was found to occur predominantly in the X (lateral) dimension. Neither brain shift nor pneumocephalus were found to be associated with lower accuracy. Finally, electrode accuracy did not impact upon the total VAT able to be generated, nor on any one specific clinical outcome.

Lower accuracy in the second electrode implantation is a well-known issue. Recognised methods to address this include adding a specific offset to the final frame co-ordinates [29] and performing staged surgery. It could also be argued that awake surgery with macrostimulation or microelectrode recordings, potentially with multiple tracts, would allow feedback of the ideal location. Other proposals include continuous irrigation after durotomy to minimise brain shift and pneumocephalus. Device hardware developments, for example local field potential recording of the best target or using directional stimulation, may facilitate compensation for lower accuracy. Nevertheless, with the lack of correspondence between accuracy and clinical outcomes or the VAT that was able to be generated, it would appear wise to not attempt elaborate methods of compensation for what in practice is apparently satisfactory accuracy.

Historically, accuracy has been appraised by comparing electrode implantations with that planned, typically using AC-PC co-ordinates. However, few consistent factors have emerged to guide improved accuracy. Furthermore, this method does not easily facilitate group analysis of multiple electrodes, comparison with functional templates (e.g. to delineate the motor subnucleus), or appraise systematic variance in targeting (i.e. electrodes could be precise and accurate but poorly planned within a nucleus). Our neuroimaging methodology addresses these shortcomings and allows a systematic appraisal of electrode inaccuracy accounting for both targeting and planning error. Using neuroimaging methods, we were able to not only identify the specific clinical situations where accuracy was lower, but appraise what factors were associated with accuracy. Our findings suggest that brain shift and pneumocephalus have less of an effect on accuracy that previously believed.

Nevertheless, a neuroimaging approach to accuracy should be seen as complimentary to rather than superseding traditional co-ordinate approaches. Strengths of the traditional co-ordinate approach include direct appraisal of how the final location compares to the intended target and comparison with individual rather than template-based anatomy. It also allows for a more clinically defined measure of accuracy in the order of millimetres from planned target, rather than the somewhat lower distances involved using our definitions of accuracy. However, when accuracy and clinical outlines are typically good, an extensive and detailed database of outcomes is necessary to identify subtle features that may be associated with accuracy in specific situations. For example, our sample size of n=38 would only be sufficient for detecting an r = 0.45 with alpha = 0.05 and beta = 0.20 prior to multiple comparisons corrections (https://sample-size.net/correlation-sample-size). With this in mind, we have not appraised the effects of age or electrode type on accuracy, for example. Notably, our methods lend themselves to easily performing these subsequent analyses, and we have freely shared our code online to do so in the hope that other larger datasets will be able to test these hypotheses in the future.

Establishing a ground truth with which to verify the accuracy of electrode localisation is an ill posed problem without using post-mortem analysis[30]. Limitations in electrode localization include the difficulty in segmenting the STN automatically at the individual level, which even with 7 Tesla MRI remains an evolving process[31]. Group templates have been introduced to obviate this issue (as well as offering additional resolution and functional segmentation)[31]. However, it is unclear how well they reflect individual anatomy, particularly in the context of a progressive neurodegenerative disease[32]. Other limitations of image processing pipelines include those related to registration (which may be ameliorated to a degree by post-operative MRI[33], and the utilisation of non-automated processes in electrode reconstruction (which is currently easier with post-operative CT and may also potentially allow directionality determination[20,34]. One must therefore be mindful that anatomical localisation data is only one aspect in considering optimal electrode targeting and must be considered alongside the complimentary neurophysiological and clinical parameters.

Overall, our accuracy was comparable with that presented in the literature, serving as a robust audit of our method (specifically, general anaesthesia throughout, frame-based, direct targeting, MRI-planned, and CT-verified)[6,7,11,35–39]. This accuracy is juxtaposed with studies of microelectrode recordings have reported revision of the original imaging-based targeting in approximately 20%. Despite this emphasis on accuracy, in our series electrode localisation did not enable prediction of clinical outcomes, in contrast to that reported in the literature[21,40,41]. One explanation for this could be a lack of statistical power related to sample size and data attrition. Furthermore, when using VAT analysis, stimulation parameters may not necessarily be optimal (due to either clinical factors or the multiple permutations of programming parameters)[42], which may obfuscate any relationship between accuracy and outcomes. Further work is required in exploring the relationship between clinical outcomes, electrode location, and determining what is clinically meaningful accuracy at the individual level.

Revision surgery has been documented as occurring in up to 15%[39,43–48] with consequent effects on healthcare services, finances, and risk of surgical complications. Myriad technologies have been proposed with the aim of improving accuracy, including the use of robotics[49–52]. In our data we identified three participants each with a single electrode location outside of the target nucleus by greater than 2mm. Reassuringly, this did not lead to any adverse neurological outcomes or unsatisfactory treatment response – and therefore neither electrode was revised – but nor was there a clear clinical indication of why discrepancy from the usual accuracy occurred. Overall, these data suggest that the accuracy achieved in routine practice is sufficient to not adversely impact upon clinical outcomes

Strengths of our study include the implementation of a relatively lightweight design and analysis strategy that integrates efficiently with a busy clinical movement disorders practice. Furthermore, we have released detailed open-source code to make this process more accessible. Limitations include the overall numbers and attrition, although these data represent a realistic reflection of what can be achieved in routine clinical practice. Improvements include focusing on streamlining data collection and optimising imaging parameters. However, the main factor that will play into improving study power will be the establishment of multi-centre collaborations and open-source datasets.

Emergence of open-source lead localisation software compliments an overall burgeoning in DBS hardware and research. Accurate appraisal of lead localisation is not only useful in deep brain stimulation surgery, but also in lesioning and cell delivery studies. Furthermore, lead localisation can be used to appraise changes to clinical practice (such as a change in imaging sequences, surgical workflow, or head position), which can now be objectively audited. This work therefore represents an ideal platform for large multi-centre audits and specifically trainee projects[53]. Such research, when performed systematically and with sufficient statistical power, may go some way to improving our understanding of accuracy and precision, as well as deriving optimal surgical workflows.

## CONCLUSIONS

In conclusion, our analysis is supportive of the accuracy in performing deep brain stimulation in a fully image-guided manner under general anaesthesia, but highlights the complexity of understanding accuracy, and cautions about lower accuracy during the second electrode. We hope that publication of these data and resources will encourage groups to utilise developments in electrode localisation, develop collaborations, and provide large open-source datasets that enhance our understanding of outcomes after deep brain stimulation.

## Data Availability

Data is not currently available according to local ethical approval.

## ACKNOWLEDGEMENTS

The authors report no acknowledgements.

## FIGURE LEGENDS

**Supplementary Figure 1: Study Flow Chart**

Flow chart of study recruitment and data attrition.

**Supplementary Figure 2: Clinical Outcomes Plotmatrix**

Plotmatrix of changes in the five clinical outcome variables. Spearman’s rank correlation coefficient (r_s_) with associated *p*-values between outcome pairs are presented in the upper triangle while raw datapoints are presented in the corresponding lower triangle. The diagonal represents differences in outcomes depending on target location (GPi or STN) with corresponding unpaired t-. GPi: Globus Pallidus Internus, LEDD: levodopa equivalent daily dose, PDQ39: Parkinson’s Disease Questionnaire, STN: Subthalamic nucleus, UPDRS: Unified Parkinson’s Disease Rating Scale

**Supplementary Figure 3: Systematic Appraisal of Electrode Accuracy and Clinical Scenarios**

I: Accuracy was not systematically different wen appraised either based on the main nucleus or motor subnucleus centre of gravity (figure 1C, left). II: Accuracy in the motor nucleus did not vary across hemispheres in the STN but did in the GP (III). A similar pattern was identified in the motor subnucleus with accuracy not varying across hemispheres in the STN (IV) but did in the GPi (V).

**Supplementary Figure 4: Systematic Appraisal of XYZ Variance in Targeting**

Raincloud plots appraising systematic variance in individual XYZ planes. Variance was identified across the group (group F-stat = 9.68, df = 7, p < 0.01) which reflected that in the X dimension of the right GPi (GPi right single-group tstat = -2.9, df = 18, p = 0.01). Distances are from the target centre of gravity, in this case the motor subnucleus.

**Supplementary Figure 5: Volume of Activated Tissue (VAT) and Clinical Outcomes**

Statistical dependencies between VAT of the main nucleus (top row) and motor subnucleus (lower row) with clinical outcomes (columns). No significant correlations or regression models were identified.

**Supplementary Figure 6: Accuracy and Clinical Outcomes**

Statistical dependencies between electrode accuracy of the main nucleus (top row) and motor subnucleus (lower row) with clinical outcomes (columns). No significant correlations or regression models were identified.

## REFERENCES

[1] Deuschl G, Schade-Brittinger C, Krack P, Volkmann J, Schäfer H, Bötzel K, et al. A Randomized Trial of Deep-Brain Stimulation for Parkinson’s Disease. New Engl J Med 2006;355:896–908. https://doi.org/10.1056/nejmoa060281.

[2] Weaver FM, Follett K, Stern M, Hur K, Harris C, Marks WJ, et al. Bilateral deep brain stimulation vs best medical therapy for patients with advanced Parkinson disease: a randomized controlled trial. Jama 2009;301:63–73. https://doi.org/10.1001/jama.2008.929.

[3] Williams A, Gill S, Varma T, Jenkinson C, Quinn N, Mitchell R, et al. Deep brain stimulation plus best medical therapy versus best medical therapy alone for advanced Parkinson’s disease (PD SURG trial): a randomised, open-label trial. Lancet Neurology 2010;9:581–91. https://doi.org/10.1016/s1474-4422(10)70093-4.

[4] Okun MS. Deep-brain stimulation for Parkinson’s disease. The New England Journal of Medicine 2012;367:1529–38. https://doi.org/10.1056/nejmct1208070.

[5] Schuepbach WMM, Rau J, Knudsen K, Volkmann J, Krack P, Timmermann L, et al. Neurostimulation for Parkinson’s Disease with Early Motor Complications. New Engl J Medicine 2013;368:610–22. https://doi.org/10.1056/nejmoa1205158.

[6] Saint-Cyr JA, Hoque T, Pereira LCM, Dostrovsky JO, Hutchison WD, Mikulis DJ, et al. Localization of clinically effective stimulating electrodes in the human subthalamic nucleus on magnetic resonance imaging. J Neurosurg 2002;97:1152–66. https://doi.org/10.3171/jns.2002.97.5.1152.

[7] McClelland S, Ford B, Senatus PB, Winfield LM, D. YE, Pullman SL, et al. Subthalamic stimulation for Parkinson disease: determination of electrode location necessary for clinical efficacy. Neurosurg Focus 2005;19:1–9. https://doi.org/10.3171/foc.2005.19.5.13.

[8] Iii SM, Vonsattel JP, Garcia RE, Amaya MD, Winfield LM, Pullman SL, et al. Relationship of clinical efficacy to postmortem-determined anatomic subthalamic stimulation in Parkinson syndrome. Clin Neuropathol 2007;26:267–75. https://doi.org/10.5414/npp26267.

[9] Pilitsis JG, Metman LV, Toleikis JR, Hughes LE, Sani SB, Bakay RAE. Factors involved in long-term efficacy of deep brain stimulation of the thalamus for essential tremor. J Neurosurg 2008;109:640–6. https://doi.org/10.3171/jns/2008/109/10/0640.

[10] Bari AA, Thum J, Babayan D, Lozano AM. Current and Expected Advances in Deep Brain Stimulation for Movement Disorders. Prog Neurol 2018;33:222–9. https://doi.org/10.1159/000481106.

[11] Burchiel KJ, McCartney S, Lee A, Raslan AM. Accuracy of deep brain stimulation electrode placement using intraoperative computed tomography without microelectrode recording. J Neurosurg 2013;119:301–6. https://doi.org/10.3171/2013.4.jns122324.

[12] Chen T, Mirzadeh Z, Ponce FA. “Asleep” Deep Brain Stimulation Surgery: A Critical Review of the Literature. World Neurosurg 2017;105:191–8. https://doi.org/10.1016/j.wneu.2017.05.042.

[13] Ho AL, Ali R, Connolly ID, Henderson JM, Dhall R, Stein SC, et al. Awake versus asleep deep brain stimulation for Parkinson’s disease: a critical comparison and meta-analysis. J Neurology Neurosurg Psychiatry 2018;89:687. https://doi.org/10.1136/jnnp-2016-314500.

[14] Ko AL, Burchiel KJ. Image-Guided, Asleep Deep Brain Stimulation. Prog Neurol 2018;33:94–106. https://doi.org/10.1159/000480984.

[15] Miocinovic S, Noecker AM, Maks CB, Butson CR, McIntyre CC. Acta Neurochirurgica Supplements. Acta Neurochir Suppl 2007;97:561–7. https://doi.org/10.1007/978-3-211-33081-4_65.

[16] D’Albis T, Haegelen C, Essert C, Fernández-Vidal S, Lalys F, Jannin P. PyDBS: an automated image processing workflow for deep brain stimulation surgery. Int J Comput Ass Rad 2015;10:117–28. https://doi.org/10.1007/s11548-014-1007-y.

[17] Horn A, Kühn AA. Lead-DBS: A toolbox for deep brain stimulation electrode localizations and visualizations. Neuroimage 2015;107:127–35. https://doi.org/10.1016/j.neuroimage.2014.12.002.

[18] Silva NM da, Rozanski VE, Cunha JPS. A 3D multimodal approach to precisely locate DBS electrodes in the basal ganglia brain region. 2015 7th Int Ieee Embs Conf Neural Eng Ner 2015:292–5. https://doi.org/10.1109/ner.2015.7146617.

[19] Lauro PM, Vanegas□Arroyave N, Huang L, Taylor PA, Zaghloul KA, Lungu C, et al. DBSproc: An open source process for DBS electrode localization and tractographic analysis. Hum Brain Mapp 2016;37:422–33. https://doi.org/10.1002/hbm.23039.

[20] Husch A, Petersen MV, Gemmar P, Goncalves J, Hertel F. PaCER - A fully automated method for electrode trajectory and contact reconstruction in deep brain stimulation. YNICL 2018;17:80–9. https://doi.org/10.1016/j.nicl.2017.10.004.

[21] Horn A, Li N, Dembek TA, Kappel A, Boulay C, Ewert S, et al. Lead-DBS v2: Towards a comprehensive pipeline for deep brain stimulation imaging. NeuroImage 2019;184:293– 316. https://doi.org/10.1016/j.neuroimage.2018.08.068.

[22] Board NC. Clinical Commissioning Policy: Deep Brain Stimulation (DBS) In Movement Disorders. 2013.

[23] (NICE) NI for H and CE. NICE Guidelines NG71: Parkinson’s disease in adults: diagnosis and management. NICE; 2017.

[24] Avants BB, Avants B, Tustison N, Johnson H, Yushkevich P. Advanced Normalization Tools: V1. 0. Insight …; 2009.

[25] Tustison NJ, Avants BB, Cook PA. The ANTs cortical thickness processing pipeline. SPIE Medical … 2013;8672:86720K – 2. https://doi.org/10.1117/12.2007128.

[26] Schönecker T, Kupsch A, Kühn AA, Schneider G-H, Hoffmann K-T. Automated Optimization of Subcortical Cerebral MR Imaging−Atlas Coregistration for Improved Postoperative Electrode Localization in Deep Brain Stimulation. Am J Neuroradiol 2009;30:1914–21. https://doi.org/10.3174/ajnr.a1741.

[27] Ewert S, Plettig P, Li N, Chakravarty MM, Collins DL, Herrington TM, et al. Toward defining deep brain stimulation targets in MNI space: A subcortical atlas based on multimodal MRI, histology and structural connectivity. Neuroimage 2018;170:271–82. https://doi.org/10.1016/j.neuroimage.2017.05.015.

[28] Allen M, Poggiali D, Whitaker K, Marshall TR, Kievit RA. Raincloud plots: a multi-platform tool for robust data visualization. Wellcome Open Res 2019;4:63. https://doi.org/10.12688/wellcomeopenres.15191.1.

[29] Park S-C, Lee JK, Kim SM, Choi EJ, Lee CS. Systematic Stereotactic Error Reduction Using a Calibration Technique in Single-Brain-Pass and Multitrack Deep Brain Stimulations. Operative Neurosurg Hagerstown Md 2018;15:72–80. https://doi.org/10.1093/ons/opx183.

[30] Reddy GD, Lozano AM. Postmortem studies of deep brain stimulation for Parkinson’s disease: a systematic review of the literature. Cell Tissue Res 2018;373:287–95. https://doi.org/10.1007/s00441-017-2672-2.

[31] Visser E, Keuken MC, Forstmann BU, Jenkinson M. Automated segmentation of the substantia nigra, subthalamic nucleus and red nucleus in 7T data at young and old age. Neuroimage 2016;139:324–36. https://doi.org/10.1016/j.neuroimage.2016.06.039.

[32] Nowacki A, Nguyen TA-K, Tinkhauser G, Petermann K, Debove I, Wiest R, et al. Accuracy of different three-dimensional subcortical human brain atlases for DBS –lead localisation. Neuroimage Clin 2018;20:868–74. https://doi.org/10.1016/j.nicl.2018.09.030.

[33] Zrinzo L, Hariz M, Hyam JA, Foltynie T, Limousin P. Letter to the Editor: A paradigm shift toward MRI-guided and MRI-verified DBS surgery. J Neurosurg 2016;124:1135–8. https://doi.org/10.3171/2015.9.jns152061.

[34] Hellerbach A, Dembek TA, Hoevels M, Holz JA, Gierich A, Luyken K, et al. DiODe: Directional Orientation Detection of Segmented Deep Brain Stimulation Leads: A Sequential Algorithm Based on CT Imaging. Stereot Funct Neuros 2018;96:335–41. https://doi.org/10.1159/000494738.

[35] Hamid NA, Mitchell RD, Mocroft P, Westby GWM, Milner J, Pall H. Targeting the subthalamic nucleus for deep brain stimulation: technical approach and fusion of pre- and postoperative MR images to define accuracy of lead placement. Journal of Neurology, Neurosurgery & Psychiatry 2005;76:409–14. https://doi.org/10.1136/jnnp.2003.032029.

[36] Bjartmarz H, Rehncrona S. Comparison of accuracy and precision between frame-based and frameless stereotactic navigation for deep brain stimulation electrode implantation. Stereotactic and Functional Neurosurgery 2007;85:235–42. https://doi.org/10.1159/000103262.

[37] Foltynie T, Zrinzo L, Martinez-Torres I, Tripoliti E, Petersen E, Holl E, et al. MRI-guided STN DBS in Parkinson’s disease without microelectrode recording: efficacy and safety. J Neurology Neurosurg Psychiatry 2011;82:358. https://doi.org/10.1136/jnnp.2010.205542.

[38] Ko AL, Ibrahim A, Magown P, Macallum R, Burchiel KJ. Factors Affecting Stereotactic Accuracy in Image-Guided Deep Brain Stimulator Electrode Placement. Stereotactic and Functional Neurosurgery 2017;95:315–24. https://doi.org/10.1159/000479527.

[39] Frizon LA, Nagel SJ, May FJ, Shao J, Maldonado-Naranjo AL, Fernandez HH, et al. Outcomes following deep brain stimulation lead revision or reimplantation for Parkinson’s disease. J Neurosurg 2018:1–6. https://doi.org/10.3171/2018.1.jns171660.

[40] Bot M, Schuurman PR, Odekerken VJJ, Verhagen R, Contarino FM, Bie RMAD, et al. Deep brain stimulation for Parkinson’s disease: defining the optimal location within the subthalamic nucleus. J Neurology Neurosurg Psychiatry 2018;89:493–8. https://doi.org/10.1136/jnnp-2017-316907.

[41] Horn A, Wenzel G, Irmen F, Huebl J, Li N, Neumann W-J, et al. Deep brain stimulation induced normalization of the human functional connectome in Parkinson’s disease. Brain 2019;142:3129–43. https://doi.org/10.1093/brain/awz239.

[42] Koeglsperger T, Palleis C, Hell F, Mehrkens JH, Bötzel K. Deep Brain Stimulation Programming for Movement Disorders: Current Concepts and Evidence-Based Strategies. Front Neurol 2019;10:410. https://doi.org/10.3389/fneur.2019.00410.

[43] Ellis T-M, Foote KD, Fernandez HH, Sudhyadhom A, Rodriguez RL, Zeilman P, et al. Reoperation for suboptimal outcomes after deep brain stimulation surgery. Neurosurgery 2008;63:754–61. https://doi.org/10.1227/01.neu.0000325492.58799.35.

[44] Falowski SM, Ooi YC, Bakay RAE. Long□Term Evaluation of Changes in Operative Technique and Hardware□Related Complications With Deep Brain Stimulation. Neuromodulation Technology Neural Interface 2015;18:670–7. https://doi.org/10.1111/ner.12335.

[45] Patel DM, Walker HC, Brooks R, Omar N, Ditty B, Guthrie BL. Adverse events associated with deep brain stimulation for movement disorders: analysis of 510 consecutive cases. Neurosurgery 2015;11 Suppl 2:190–9. https://doi.org/10.1227/neu.0000000000000659.

[46] Falowski SM, Bakay RAE. Revision Surgery of Deep Brain Stimulation Leads. Neuromodulation Technology Neural Interface 2016;19:443–50. https://doi.org/10.1111/ner.12404.

[47] Rolston JD, Englot DJ, Starr PA, Larson PS. An unexpectedly high rate of revisions and removals in deep brain stimulation surgery: Analysis of multiple databases. Parkinsonism Relat D 2016;33:72–7. https://doi.org/10.1016/j.parkreldis.2016.09.014.

[48] Nickl RC, Reich MM, Pozzi NG, Fricke P, Lange F, Roothans J, et al. Rescuing Suboptimal Outcomes of Subthalamic Deep Brain Stimulation in Parkinson Disease by Surgical Lead Revision. Neurosurgery 2019. https://doi.org/10.1093/neuros/nyz018.

[49] Xu F, Jin H, Yang X, Sun X, Wang Y, Xu M, et al. Improved accuracy using a modified registration method of ROSA in deep brain stimulation surgery. Neurosurg Focus 2018;45:E18. https://doi.org/10.3171/2018.4.focus1815.

[50] Goia A, Gilard V, Lefaucheur R, Welter M, Maltête D, Derrey S. Accuracy of the robot□assisted procedure in deep brain stimulation. Int J Medical Robotics Comput Assisted Surg 2019;15. https://doi.org/10.1002/rcs.2032.

[51] Liu L, Mariani SG, Schlichting ED, Grand S, Lefranc M, Seigneuret E, et al. Frameless ROSA® Robot-Assisted Lead Implantation for Deep Brain Stimulation: Technique and Accuracy. Oper Neurosurg 2019. https://doi.org/10.1093/ons/opz320.

[52] VanSickle D, Volk V, Freeman P, Henry J, Baldwin M, Fitzpatrick CK. Electrode Placement Accuracy in Robot-Assisted Asleep Deep Brain Stimulation. Ann Biomed Eng 2019;47:1212–22. https://doi.org/10.1007/s10439-019-02230-3.

[53] Chari A, Jamjoom AA, Edlmann E, Ahmed AI, Coulter IC, Ma R, et al. The British Neurosurgical Trainee Research Collaborative: Five years on. Acta Neurochir 2017;160:23– 8. https://doi.org/10.1007/s00701-017-3351-5.

