## Supplementary figures and images for "Increased variance in second electrode accuracy during deep brain stimulation and its relationship to pneumocephalus, brain shift, and clinical outcomes: a retrospective cohort study"

### supplemental figure 1

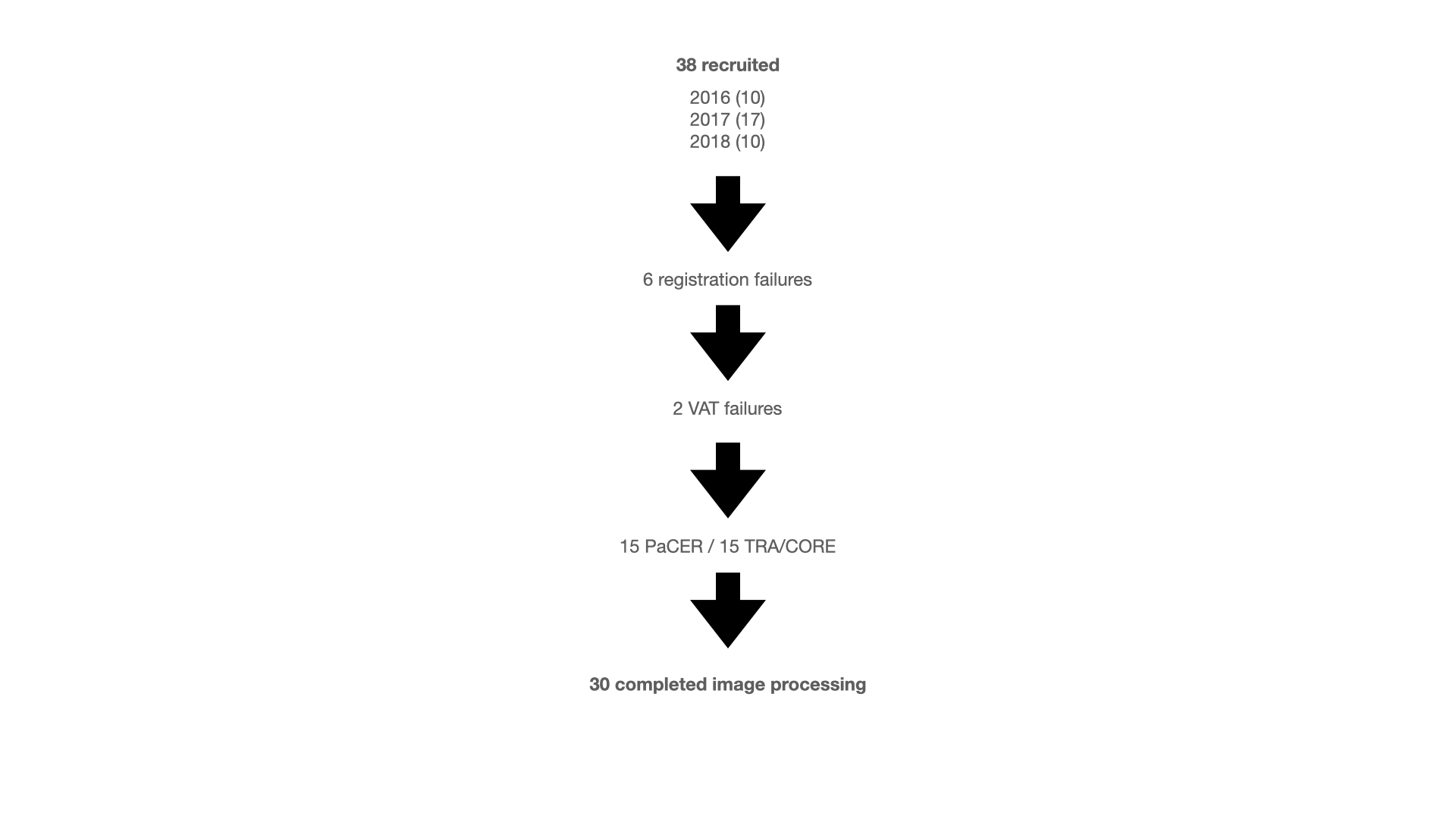

### supplemental figure 2

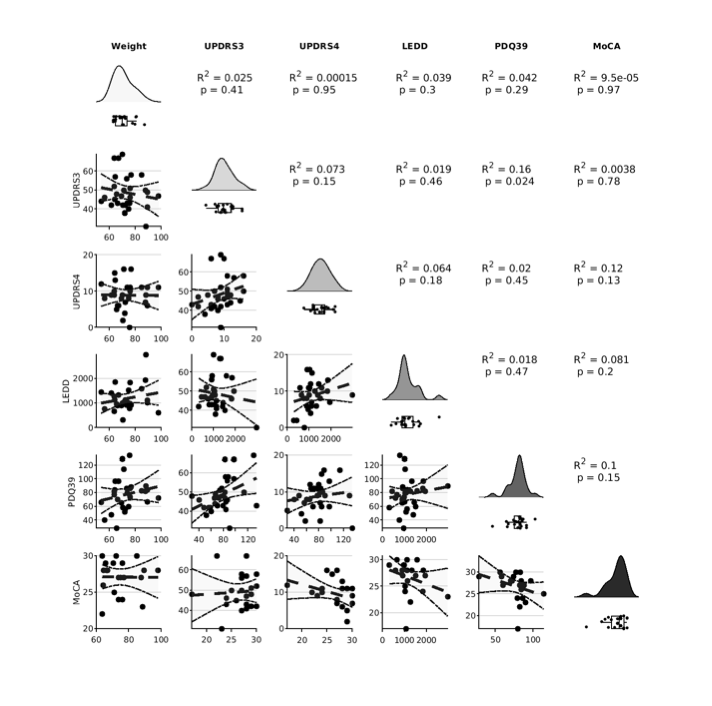

### supplemental figure 3

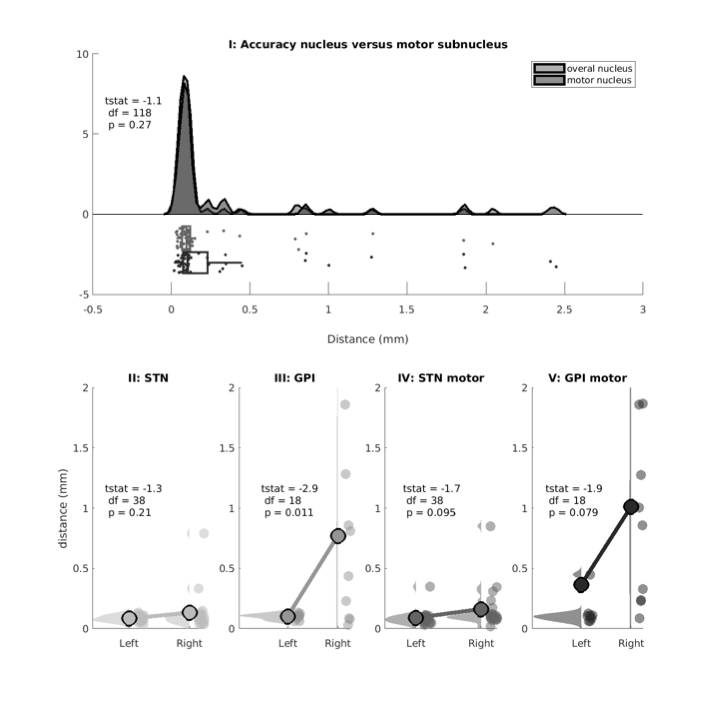

### supplemental figure 4

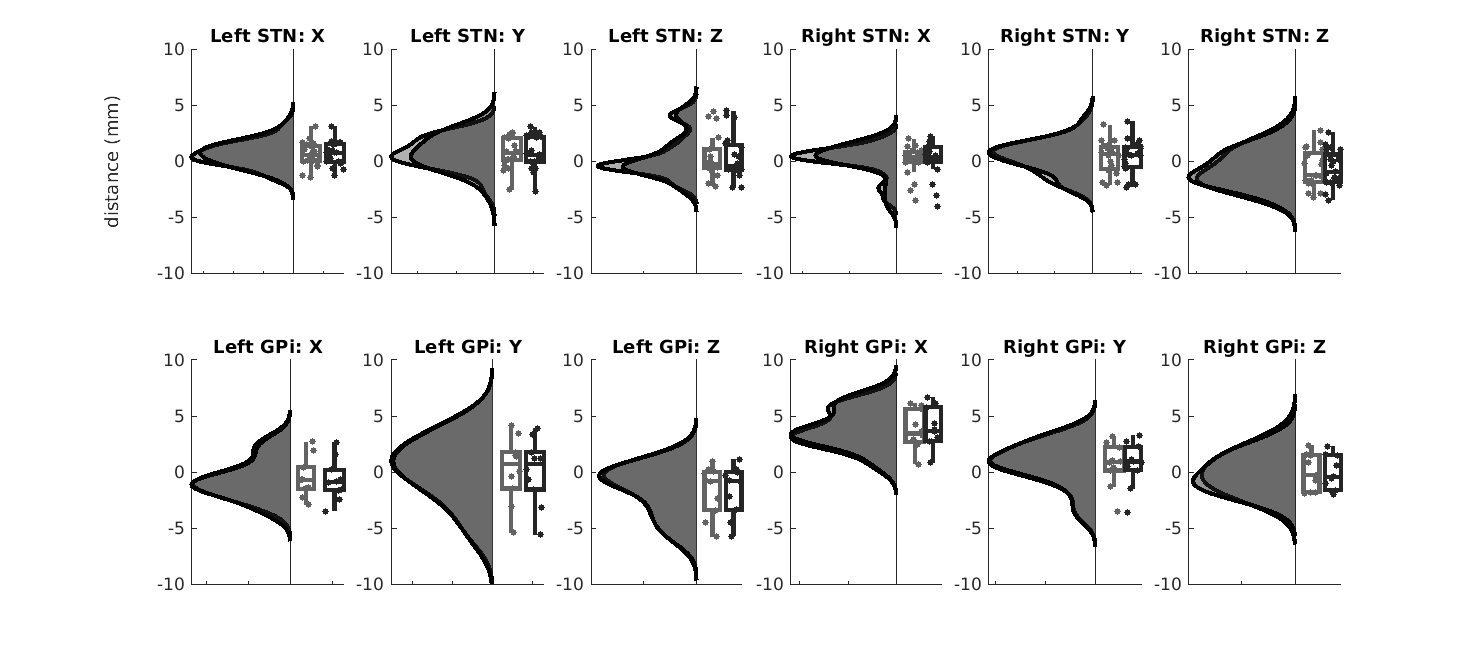

### supplemental figure 5

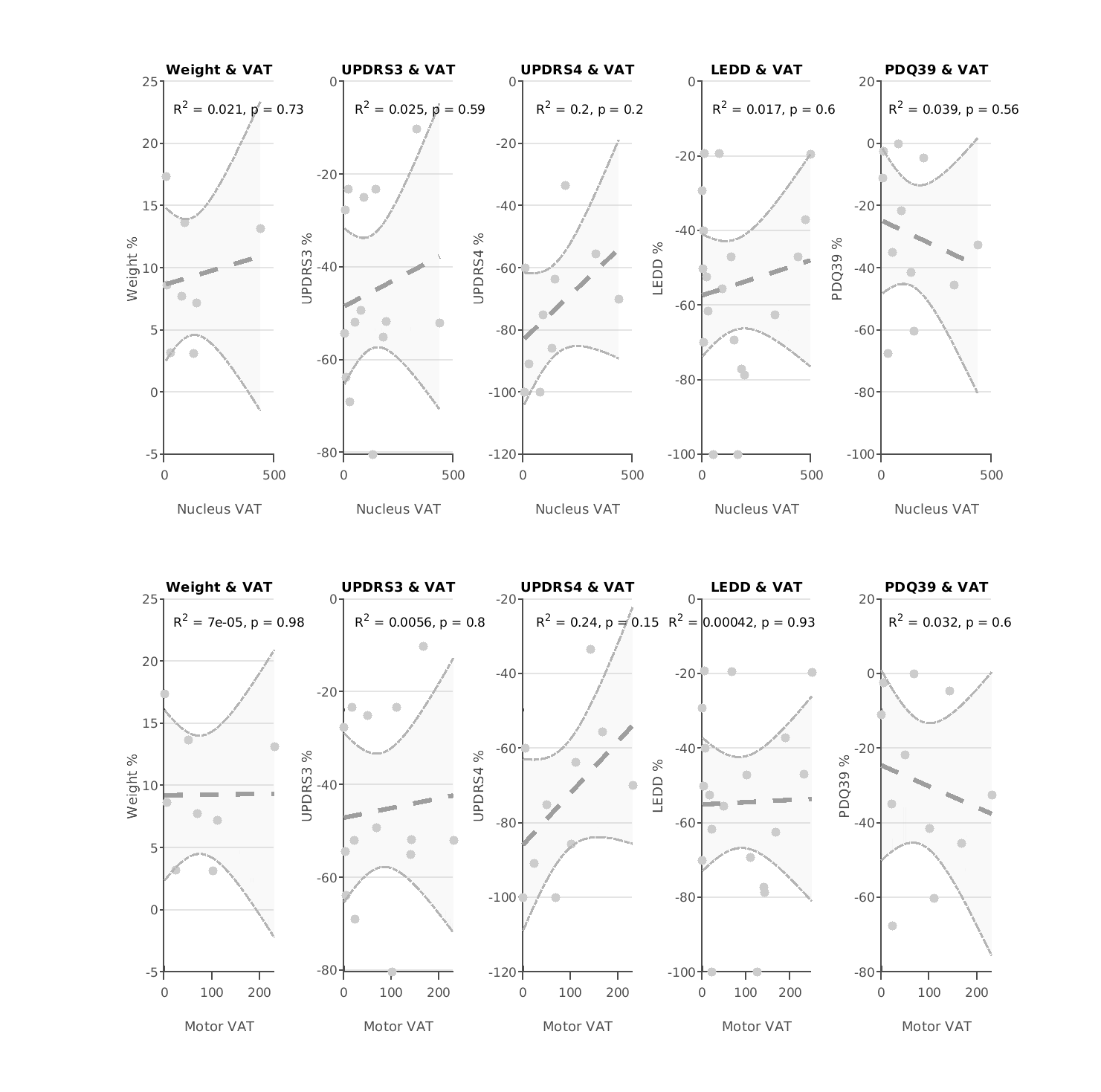

### supplemental figure 6

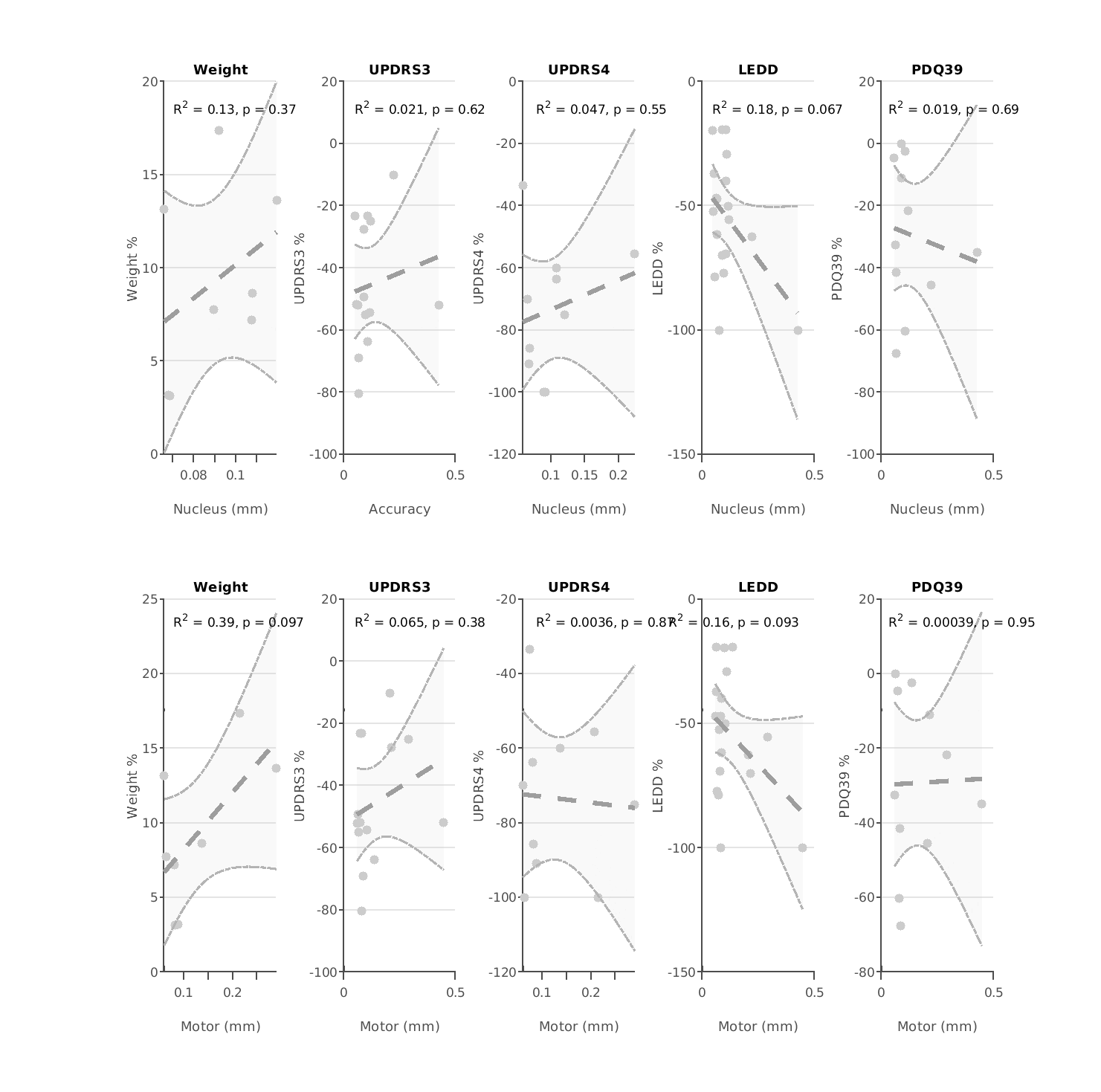
